# NeoCLIP: A Self-Supervised Foundation Model for the Interpretation of Neonatal Radiographs

**DOI:** 10.1101/2024.12.03.24318410

**Authors:** Yixuan Huang, Puneet Sharma, Anil Palepu, Nathaniel Greenbaum, Andrew Beam, Kristyn Beam

## Abstract

**Importance:** Artificial intelligence (AI) based on deep learning has shown promise in adult and pediatric populations in the interpretation of medical imaging to make important diagnostic and management recommendations. However, there has been little work developing new AI methods for neonatal populations.

**Objective:** To develop a novel, deep contrastive learning model to predict a comprehensive set of pathologies from radiographs relevant to neonatal intensive care.

**Design, Setting, and Participants:** We identified a retrospective cohort of infants who obtained a radiograph while admitted to a large neonatal intensive care unit in Boston, MA from January 2008 to December 2023. After collecting radiographs with corresponding reports and relevant demographics for all subjects, we randomized the cohort into three sets: training (80%), validation (10%), and test (10%).

**Interventions:** We developed a deep learning model, NeoCLIP, to identify 15 unique pathologies and 5 medical devices relevant to neonatal intensive care on plain film radiographs. The pathologies were automatically extracted from radiology reports using a custom pipeline based on large language models.

**Main Outcomes and Measures:** We compared the performance of our model, as defined by AUROC, against various baseline methods.

**Results:** We identified 4,629 infants which were randomized into the training (3,731 infants), validation (419 infants), and test (479 infants) sets. In total, we collected 20,154 radiographs with a corresponding 15,795 reports. The AUROC of our model was greater than all baseline methods for every radiographic finding other than portal venous gas. The addition of demographics improved the AUROC of our model for all findings, but the difference was not statistically significant.

**Conclusions and Relevance:** NeoCLIP successfully identified a broad set of pathologies and medical devices on neonatal radiographs, outperforming similar models developed for adult populations. This represents the first such application of advanced AI methodologies to interpret neonatal radiographs.

## Introduction

In the past few decades, there has been a major transition in the way medicine is practiced. A discipline that once relied on paper charts and radiographs has become fully electronic. With the advent of the electronic medical record and digitization of medical imaging, there is a profound amount of data being generated by healthcare. It has been estimated that thirty percent of the entire world’s data volume is generated by the healthcare industry (1). In this data, there is an abundance of information that could improve the care of patients; however, the volume is becoming far too large for human processing. Fortunately, there has been substantial progress made in recent years in artificial intelligence (AI), largely driven by advancements in deep learning (2). The advancements in this technology have made it possible to learn from this data to improve clinical care and further improve the way medicine is practiced.

In the field of radiology, deep learning has allowed for the interpretation of medical imaging to make important diagnostic and management recommendations (3-5). In particular, vision-language contrastive learning models trained on paired radiographs and reports have shown promise (6-10). These models typically leverage general-purpose foundation models that are initially trained on large-scale datasets and subsequently adapted to domain-specific contexts and knowledge. In this framework, models are trained to align radiographic images with their corresponding reports. The models learn, in a self-supervised manner, to identify image features that correspond to the information contained in the reports. This allows the model to associate clinical features present in the images with diagnostic descriptions without being trained on explicit labels. Although these advancements have been demonstrated in adult and pediatric populations, they have not been systematically developed for neonates. This disparity is particularly interesting given the reliance on clinical imaging in the management of critically ill neonates. Many common pathologies of neonatal-perinatal medicine are either screened for or diagnosed on plain film radiographs. It is common to screen for bronchopulmonary dysplasia (BPD) and respiratory distress syndrome (RDS) on chest x-ray (11-12). Furthermore, radiographic signs such as pneumatosis or portal venous gas are essential for the diagnosis of necrotizing enterocolitis (13). These unique neonatal pathologies are of course in addition to other important indications for radiographs in critical care such as post-procedural imaging.

A deep learning model that could interpret neonatal imaging would both be novel and potentially augment the care provided to this vulnerable population. In doing so, it could allow for faster diagnosis and interpretation of complex pathophysiologies and management decisions. Furthermore, these methodologies may serve as a gateway to improved management of these complex disease states by detecting subtle findings in imaging not appreciable to the human eye As such, the objective of our study was to develop a foundation model for neonatal imaging to interpret radiographs to identify common pathologies and findings relevant to neonatal intensive care.

## Methods

### Study Population

We conducted a retrospective study on a cohort of all infants admitted to the neonatal intensive care unit (NICU) at Beth Israel Deaconess Medical Center in Boston, MA from January 2008 to December 2023. Any infant who did not obtain a radiograph during their admission to the NICU was excluded. For all the infants in our cohort, we collected radiographs and corresponding reports from their admission to the NICU. In addition to the radiographic data, we also collected a predefined list of demographic data (**Table 1**). We used simple random sampling to then split the enrolled cohort into three distinct sets: training, validation, and test. The sampling was performed at the patient level to ensure that all radiographs and reports from a single patient remained in the same set. We used Python’s random module with a random seed of 1 for reproducibility.

**Table 1.** Randomization of Cohort.

|  | **Training cohort** |  |  | **Validation cohort** |  |  | **Test cohort** |  |  |
| --- | --- | --- | --- | --- | --- | --- | --- | --- | --- |
|  | **Patients**<br>**(n =**<br>**3,731)** | **Radiographs**<br>**(n = 16,204)** | **Reports**<br>**(n =**<br>**12,727)** | **Patients**<br>**(n =**<br>**419)** | **Radiographs**<br>**(n = 1,915)** | **Reports**<br>**(n =**<br>**1,454)** | **Patients**<br>**(n =**<br>**479)** | **Radiographs**<br>**(n = 2,035)** | **Reports**<br>**(n =**<br>**1,614)** |
| **Sex (%)** |  |  |  |  |  |  |  |  |  |
| **Female** | 1,581<br>(42) | 7,261 (45) | 5,716<br>(45) | 241 (58) | 769 (40) | 611 (42) | 265 (55) | 886 (44) | 702 (43) |
| **Male** | 2,150<br>(58) | 8,943 (55) | 7,011<br>(55) | 178 (42) | 1,146 (60) | 843 (58) | 214 (45) | 1,149 (56) | 912 (57) |
| **Race and Ethnicity of**<br>**Mother (%)** |  |  |  |  |  |  |  |  |  |
| **Asian** | 403 (11) | 1,565 (10) | 1,258<br>(10) | 43 (10) | 246 (13) | 183 (13) | 37 (8) | 230 (11) | 185 (11) |
| **Black or African**<br>**American** | 522 (14) | 2,832 (17) | 2,202<br>(17) | 67 (16) | 412 (22) | 318 (22) | 63 (13) | 334 (16) | 269 (17) |
| **Hispanic** | 649 (17) | 2,872 (18) | 2,213<br>(17) | 77 (18) | 428 (22) | 320 (22) | 84 (18) | 421 (21) | 317 (20) |
| **White** | 2,143<br>(57) | 8,900 (54) | 7,024<br>(55) | 232 (56) | 829 (43) | 633 (44) | 294 (60) | 1,049 (51) | 842 (52) |
| **Other** | 14 (1) | 35 (1) | 30 (1) | 0 (0) | 0 (0) | 0 (0) | 1 (1) | 1 (1) | 1 (1) |
| **Birth weight (g), mean (SD)** | 2,405 (1,004) |  |  | 2,379 (1,005) |  |  | 2,408 (1,004) |  |  |
| **Gestational age (weeks), mean (SD)** | 34.4 (4.3) |  |  | 34.4 (4.3) |  |  | 34.5 (4.2) |  |  |
| **Positive cases (%)** |  |  |  |  |  |  |  |  |  |
| **Atelectasis** |  | 5,504 (34) | 4,459 (35) |  | 662 (35) | 514 (35) |  | 740 (36) | 614 (38) |
| **Intestinal Atresia** |  | 138 (1) | 125 (1) |  | 27 (1) | 19 (1) |  | 25 (1) | 21 (1) |
| **Bronchopulmonary Dysplasia** |  | 1,759 (11) | 1,528 (12) |  | 170 (9) | 144 (10) |  | 323 (16) | 288 (18) |
| **Cardiomegaly** |  | 971 (6) | 781 (6) |  | 125 (7) | 95 (7) |  | 104 (5) | 82 (5) |
| **Endotracheal Tube Placement** |  | 6,764 (42) | 5,255 (41) |  | 739 (39) | 564 (39) |  | 911 (45) | 729 (45) |
| **Meconium Aspiration Syndrome** |  | 377 (2) | 299 (2) |  | 46 (2) | 36 (2) |  | 31 (2) | 27 (2) |
| **Nasogastric Tube Placement** |  | 7,926 (49) | 6,419 (50) |  | 970 (51) | 779 (54) |  | 987 (49) | 826 (51) |
| **Intestinal Obstruction** |  | 368 (2) | 315 (2) |  | 71 (4) | 51 (4) |  | 74 (3) | 55 (3) |
| **Pulmonary Edema** |  | 709 (4) | 611 (5) |  | 59 (3) | 48 (3) |  | 62 (3) | 48 (3) |
| **Peripherally Inserted**<br>**Central Catheter**<br>**Placement** |  | 2,278 (14) | 1,597<br>(13) |  | 281 (15) | 193<br>(13) |  | 283 (14) | 218<br>(14) |
| **Pleural Effusion** |  | 751 (5) | 636 (5) |  | 93 (5) | 74 (5) |  | 79 (4) | 61 (4) |
| **Pneumonia** |  | 236 (1) | 208 (2) |  | 27 (1) | 24 (2) |  | 28 (1) | 25 (2) |
| **Pneumatosis Intestinalis** |  | 489 (3) | 391 (3) |  | 54 (3) | 41 (3) |  | 53 (3) | 43 (3) |
| **Pneumothorax** |  | 924 (6) | 726 (6) |  | 71 (4) | 59 (4) |  | 72 (4) | 60 (4) |
| **Pneumoperitoneum** |  | 95 (1) | 76 (1) |  | 21 (1) | 13 (1) |  | 13 (1) | 12 (1) |
| **Portal Venous Gas** |  | 208 (1) | 136 (1) |  | 11 (1) | 7 (0) |  | 16 (1) | 10 (1) |
| **Respiratory Distress**<br>**Syndrome** |  | 6,154 (38) | 4,661<br>(37) |  | 672 (35) | 510<br>(35) |  | 808 (40) | 605<br>(37) |
| **Transient Tachypnea**<br>**Of The Newborn** |  | 456 (3) | 402 (3) |  | 57 (3) | 53 (4) |  | 63 (3) | 54 (3) |
| **Umbilical Artery**<br>**Catheter Placement** |  | 1,717 (11) | 1,150<br>(9) |  | 226 (12) |  |  | 209 (10) | 151 (9) |
| **Umbilical Vein**<br>**Catheter Placement** |  | 5,883 (36) | 3,902<br>(31) |  | 697 (36) |  |  | 731 (36) | 478<br>(30) |

### Model Development and Pre-Training

We aimed to develop a multitasked model with the ability to identify the presence of 15 neonatal pathologies/radiographic findings and five types of medical devices. The pathologies and findings include intestinal atresia, BPD, cardiomegaly, meconium aspiration syndrome (MAS), pneumonia, RDS, transient tachypnea of the newborn (TTN), atelectasis, intestinal obstruction, pulmonary edema, pleural effusion, pneumatosis intestinalis, pneumothorax, pneumoperitoneum, and portal venous gas. The medical devices include endotracheal tube (ETT), nasogastric tube (NGT), peripherally inserted central catheter (PICC), umbilical artery catheter (UAC), and umbilical vein catheter (UVC).

We adopted the BioViL-T, a domain-specific vision-language model trained on adult chest X-rays and their associated reports (15). The vision encoder of the model is ResNet-50 (a convolutional neural network), and the text encoder is a BERT large language model (16). ResNet-50 is a deep learning model that is pre-trained on ImageNet, a large-scale imaging dataset containing millions of diverse images not specific to medicine (17). We conducted neonatal-specific pre-training of the model using the radiographs and reports. During the pre-training process, the image and text embeddings generated by the vision and text encoders were projected to a shared latent space and a similarity score was calculated for each pair of image and report. Using common contrastive language-image pre-training (CLIP) model training approaches, the models were trained to maximize the similarity scores of the true image-text pairs while minimizing the scores of other pairs (18). We trained the model for 50 epochs with a batch size of 256, using the Adam optimizer with an initial learning rate of 0.0001. To mitigate overfitting, we applied data augmentation techniques such as random rotation, horizontal and vertical flips, affine transformations, and color jittering (19-20). Additionally, to enhance training efficiency, all images were resized to 224x224 pixels. The pixel values were normalized using a mean of [0.485, 0.456, 0.406] and a standard deviation of [0.229, 0.224, 0.225].

### Large Language Model (LLM) Label Extraction

To enhance model performance, we fine-tuned the pre-trained models using supervised training techniques. This approach required labels indicating disease status and were not readily available in the dataset. To address this, we employed Mixtral-8x7B, a LLM, to extract these labels from the radiology reports, creating label-specific prompts to ask the model to identify the presence of each label in the report. For each disease and device label, two neonatologists provided definitions and descriptions which were included in the corresponding prompt. Additionally, we used “few-shot learning” in which a small set of training examples, including both positive and negative examples for each label, was provided in each prompt to improve performance. After the labels were generated, we used stratified sampling to select 10 positive and 10 negative labels for each disease, which were then reviewed by our neonatologists to determine accuracy, specificity, and sensitivity of the LLM.

### Development of NeoCLIP

We continued task-specific fine-tuning on the pre-trained model, using the labels generated by the LLM as targets. We utilized the vision encoder from the pre-trained CLIP model in the previous step, adding a multilayer perceptron and a final linear layer as a classification head. We additionally fine-tuned a version where the gestational age and birth weight of patients, standardized to zero mean and unit variance, were concatenated to the vision encoder’s output. We trained the model with a binary cross-entropy loss function to optimize the model for the detection of specific neonatal findings. This fine-tuning phase was conducted for 50 epochs, with a batch size of 256, using the Adam optimizer and a learning rate of 0.00005. We selected the final checkpoint of the model with the highest average Area Under the Receiver Operating Characteristic curve (AUROC) for the validation set.

### Model Benchmarks

We compared the performance of NeoCLIP against several standard baseline methods. This included a logistic regression model based solely on the gestational age and birth weight of the patient, the BioViL-T imaging model, and the ResNet-50 imaging model. We also assessed the reliability of our model over time by examining whether the chronologic age of the infants at time of radiograph impacted its performance. To examine the features the model used for prediction, we generated saliency maps for the best and worst performing labels. These maps highlight regions of a given input image that are important for producing the prediction on that image and therefore lend insight into how the model performs its task.

## Results

### Study Cohort

We identified 7,986 infants who were admitted to the NICU at Beth Israel Deaconess Medical Center during our enrollment period. Of these infants, 3,357 were excluded as they did not have a radiograph obtained during their admission. This resulted in a cohort of 4,629 infants for whom we obtained demographic data and all radiographs with corresponding reports. In total, we obtained 20,154 radiographs with a corresponding 15,795 reports. The breakdown of radiograph type in our cohort was: 10,793 chest x-rays, 3,413 abdominal x-rays, and 5,948 babygrams (chest and abdomen in one film). The reason for the discrepancy between radiographs and reports is that often one report described multiple radiographs, such as when multiple images were collected in succession. We then randomized the cohort into the three sets noted previously. The distribution after randomization is as follows:

1. Training set: 3,731 patients (80%) with 16,204 images and 12,727 reports.
2. Validation set: 419 patients (10%) with 1,915 images and 1,454 reports.
3. Test set: 479 (10%) with 2,035 images and 1,614 reports.

The mean gestational age and weight of all three sets were similar after randomization - approximately 34 weeks and 2.4 kg respectively. All other relevant demographic data and summary of images collected are summarized in **Table 1**.

### Label Extraction Performance

The performance of our LLM-based label extraction was assessed by two neonatologists on a stratified sample of 10 positive and 10 negative cases for each label. The kappa statistic measuring interrater reliability between the two neonatologists was 0.81 (95% CI, 0.72 - 0.91). The overall accuracy of the LLM-extracted labels was 0.90, with perfect accuracy noted for seven labels. The sensitivity and specificity of the labels were 0.98 and 0.84, respectively. **Table 2** summarizes the accuracy, sensitivity, and specificity for all 20 labels.

**Table 2.** Performance of Large Language Model in Label Extraction.

| **Label** | **Accuracy** | **Sensitivity** | **Specificity** |
| --- | --- | --- | --- |
| **Atelectasis** | **0.90** | **0.90** | **0.90** |
| **Intestinal Atresia** | **0.60** | **1.00** | **0.56** |
| **Bronchopulmonary Dysplasia** | **1.00** | **1.00** | **1.00** |
| **Cardiomegaly** | **0.95** | **1.00** | **0.91** |
| **Endotracheal Tube Placement** | **1.00** | **1.00** | **1.00** |
| **Meconium Aspiration Syndrome** | **0.85** | **1.00** | **0.77** |
| **Nasogastric Tube Placement** | **0.75** | **0.86** | **0.69** |
| **Intestinal Obstruction** | **0.65** | **1.00** | **0.59** |
| **Pulmonary Edema** | **1.00** | **1.00** | **1.00** |
| **Peripherally Inserted Central Catheter Placement** | **0.95** | **1.00** | **0.91** |
| **Pleural Effusion** | **0.90** | **0.9** | **0.9** |
| **Pneumonia** | **0.80** | **1.00** | **0.71** |
| **Pneumatosis Intestinalis** | **1.00** | **1.00** | **1.00** |
| **Pneumothorax** | **0.95** | **1.00** | **0.91** |
| **Pneumoperitoneum** | **0.85** | **1.00** | **0.77** |
| **Portal Venous Gas** | **1.00** | **1.00** | **1.00** |
| **Respiratory Distress Syndrome** | **0.85** | **1.00** | **0.77** |
| **Transient Tachypnea of the Newborn** | **1.00** | **1.00** | **1.00** |
| **Umbilical Artery Catheter Placement** | **0.95** | **1.00** | **0.91** |
| **Umbilical Vein Catheter Placement** | **1.00** | **1.00** | **1.00** |
| **Total** | **0.90** | **0.98** | **0.84** |

### Model Performance

After NeoCLIP was pre-trained and fine-tuned, we assessed its performance on the test set. For each of the twenty tasks, we determined the AUROC, which is summarized in **Table 3**. For the pathology labels, the model performed best on BPD (0.96, SD = 0.01) and worst on pulmonary edema (0.65, SD = 0.04). The model performed better on medical device labels with its greatest AUROCs on ETT (0.95, SD = 0.01) and UVC placement (0.97, SD = 0.00). Our models outperform both the logistic regression and base ResNet-50 models on all labels, except intestinal atresia (0.81, SD = 0.04 v 0.83, SD = 0.04) and MAS (0.90, SD = 0.04 v 0.90, SD = 0.04), but the differences are not statistically significant. Similarly, our model outperforms both versions of the BioViL-T model on all labels other than portal venous gas (0.73, SD = 0.06 v 0.79, SD = 0.03), with the difference on this label also remaining statistically insignificant. As described, our imaging model performs well, but it often performs even better with the addition of the patient’s gestational age and birthweight. As noted in **Table 3**, the AUROC of 12 labels improve with the addition of these important demographic details. Among these, ETT placement and pneumoperitoneum had statistically significant improvements with the addition of gestational age and birthweight.

**Table 3.** Performance of Our Model Compared to Multiple Controls.

|  | Logistic regression on<br>gestational age and<br>birth weight | ResNet-50 | BioViL-T | BioViL-T with<br>gestational age and<br>birth weight | NeoCLIP | NeoCLIP with<br>gestational age and<br>birth weight |
| --- | --- | --- | --- | --- | --- | --- |
| **Atelectasis (SD)** | 0.596 (0.013) | 0.682<br>(0.012) | 0.680<br>(0.013) | 0.681 (0.012) | **0.700**<br>**(0.012)** | 0.697 (0.012) |
| **Intestinal Atresia (SD)\*** | 0.520 (0.065) | **0.834**<br>**(0.042)** | 0.823<br>(0.045) | 0.806 (0.043) | 0.800<br>(0.048) | 0.810 (0.039) |
| **Bronchopulmonary Dysplasia**<br>**(SD)** | 0.834 (0.009) | 0.941<br>(0.007) | 0.942<br>(0.006) | 0.956 (0.005) | 0.958<br>(0.005) | **0.966 (0.005)** |
| **Cardiomegaly (SD)\*** | 0.588 (0.029) | 0.652<br>(0.028) | 0.647<br>(0.028) | 0.665 (0.028) | **0.757**<br>**(0.022)** | 0.734 (0.025) |
| **Endotracheal Tube Placement (SD)** | 0.680 (0.012) | 0.833<br>(0.009) | 0.834<br>(0.009) | 0.855 (0.008) | 0.954<br>(0.005) | **0.955 (0.004)** |
| **Meconium Aspiration Syndrome (SD)\*** | **0.900 (0.036)** | 0.830<br>(0.036) | 0.826<br>(0.036) | 0.885 (0.038) | 0.849<br>(0.033) | 0.896 (0.036) |
| **Nasogastric Tube Placement (SD)** | 0.692 (0.012) | 0.777<br>(0.010) | 0.775<br>(0.010) | 0.781 (0.010) | 0.863<br>(0.008) | **0.868 (0.008)** |
| **Intestinal Obstruction (SD)** | 0.712 (0.028) | 0.868<br>(0.022) | 0.867<br>(0.022) | 0.875 (0.021) | 0.881<br>(0.019) | **0.886 (0.017)** |
| **Pulmonary Edema (SD)** | 0.611 (0.036) | 0.536<br>(0.040) | 0.536<br>(0.040) | 0.607 (0.038) | 0.649<br>(0.037) | **0.650 (0.035)** |
| **Peripherally Inserted Central Catheter Placement (SD)** | 0.734 (0.014) | 0.762<br>(0.016) | 0.762<br>(0.016) | 0.789 (0.015) | 0.819<br>(0.013) | **0.823 (0.013)** |
| **Pleural Effusion (SD)** | 0.746 (0.028) | 0.775<br>(0.024) | 0.777<br>(0.024) | 0.783 (0.026) | 0.778<br>(0.025) | **0.789 (0.024)** |
| **Pneumonia (SD)** | 0.580 (0.059) | 0.666<br>(0.057) | 0.684<br>(0.056) | 0.646 (0.063) | **0.768**<br>**(0.051)** | 0.746 (0.057) |
| **Pneumatosis Intestinalis (SD)** | 0.589 (0.024) | 0.860<br>(0.028) | 0.858<br>(0.028) | 0.862 (0.027) | **0.893**<br>**(0.019)** | 0.892 (0.020) |
| **Pneumothorax (SD)** | 0.669 (0.029) | 0.802<br>(0.025) | 0.793<br>(0.025) | 0.777 (0.023) | **0.879**<br>**(0.019)** | 0.870 (0.020) |
| **Pneumoperitoneum (SD)** | 0.686 (0.061) | 0.686<br>(0.091) | 0.681<br>(0.090) | 0.762 (0.071) | 0.814<br>(0.054) | **0.838 (0.061)** |
| **Portal Venous Gas (SD)** | 0.558 (0.073) | 0.759<br>(0.058) | 0.826<br>(0.028) | **0.788 (0.026)** | 0.734<br>(0.055) | 0.641 (0.059) |
| **Respiratory Distress Syndrome (SD)** | 0.579 (0.012) | 0.808<br>(0.010) | 0.808<br>(0.009) | 0.845 (0.009) | 0.865<br>(0.008) | **0.876 (0.007)** |
| **Transient Tachypnea of the Newborn (SD)** | 0.851 (0.014) | 0.841<br>(0.020) | 0.841<br>(0.020) | 0.881 (0.014) | 0.858<br>(0.023) | **0.890 (0.015)** |
| **Umbilical Artery Catheter Placement (SD)** | 0.645 (0.022) | 0.890<br>(0.014) | 0.892<br>(0.012) | 0.916 (0.011) | 0.940<br>(0.010) | **0.948 (0.010)** |
| **Umbilical Vein Catheter Placement (SD)** | 0.558 (0.014) | 0.921<br>(0.006) | 0.923<br>(0.006) | 0.931 (0.006) | 0.967<br>(0.004) | **0.968 (0.004)** |

We also assessed the stability of NeoCLIP’s performance by assessing the AUROCs for all labels as a function of time the image was obtained. **Figure 1** summarizes the model’s performance based on the week the image was obtained during the patient’s admission. For the labels that had images obtained each week, the AUROCs remained relatively stable. Notable exceptions include pneumonia, which had an AUROC increase after week 4 and UVC placement which had an AUROC decrease at week 2. We also assessed the model’s performance during the first week of life as a function of day of life the image was obtained (**Figure 2**). As with the case with the weekly data points, not every label had images obtained each day during the first week of life. However, for those labels that did, we see a similar stability with the only major exception being pneumothorax which sees a large decline in AUROC on day 3. In addition to these stability assessments, we also generated saliency maps to better explain our model’s performance. **Figure 3** includes sample maps for the five best and worst performing models as defined by AUROCs.

**Figure 1.**
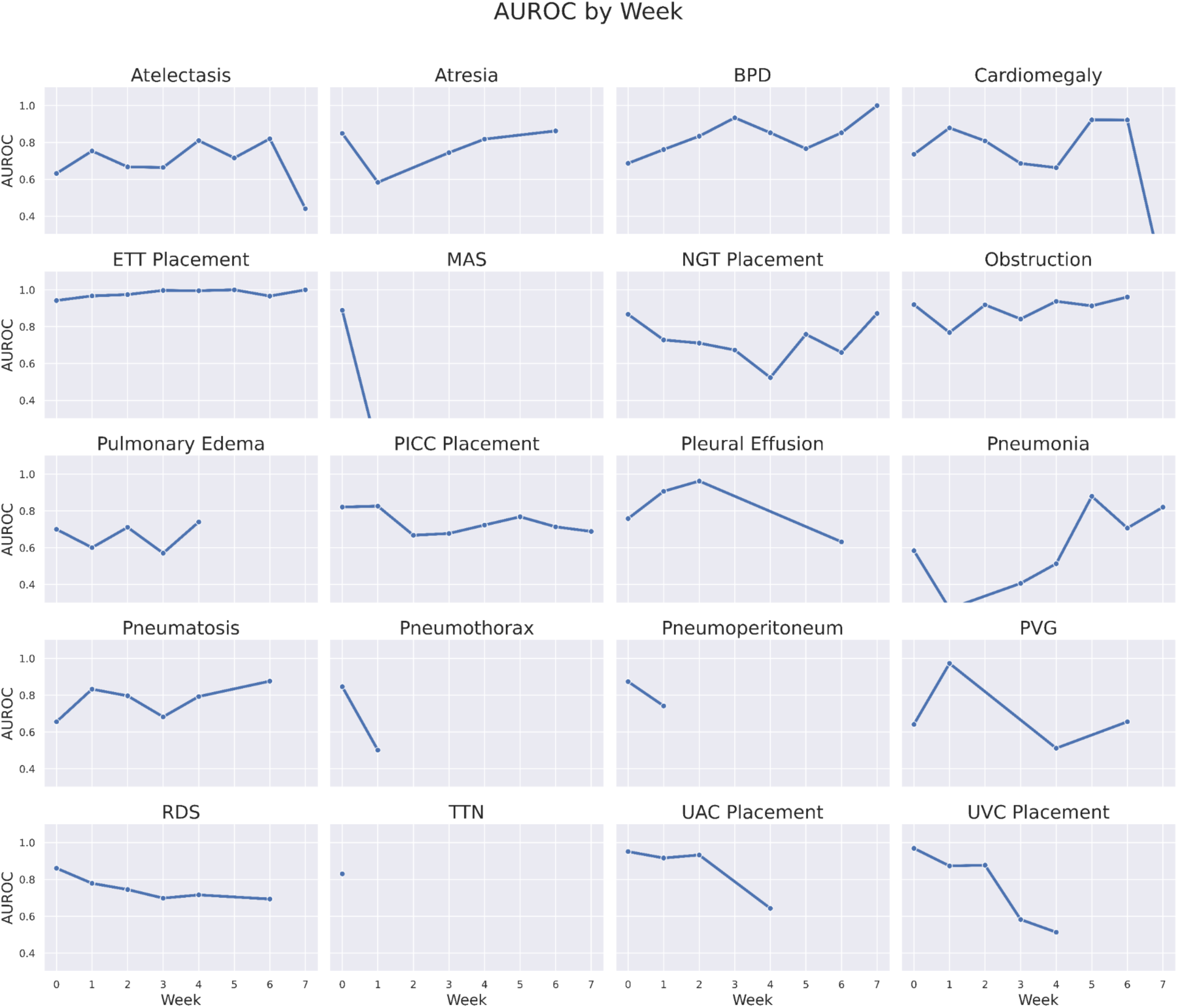
AUROC of Our Model by the Week Image Was Obtained During Admission.

**Figure 2.**
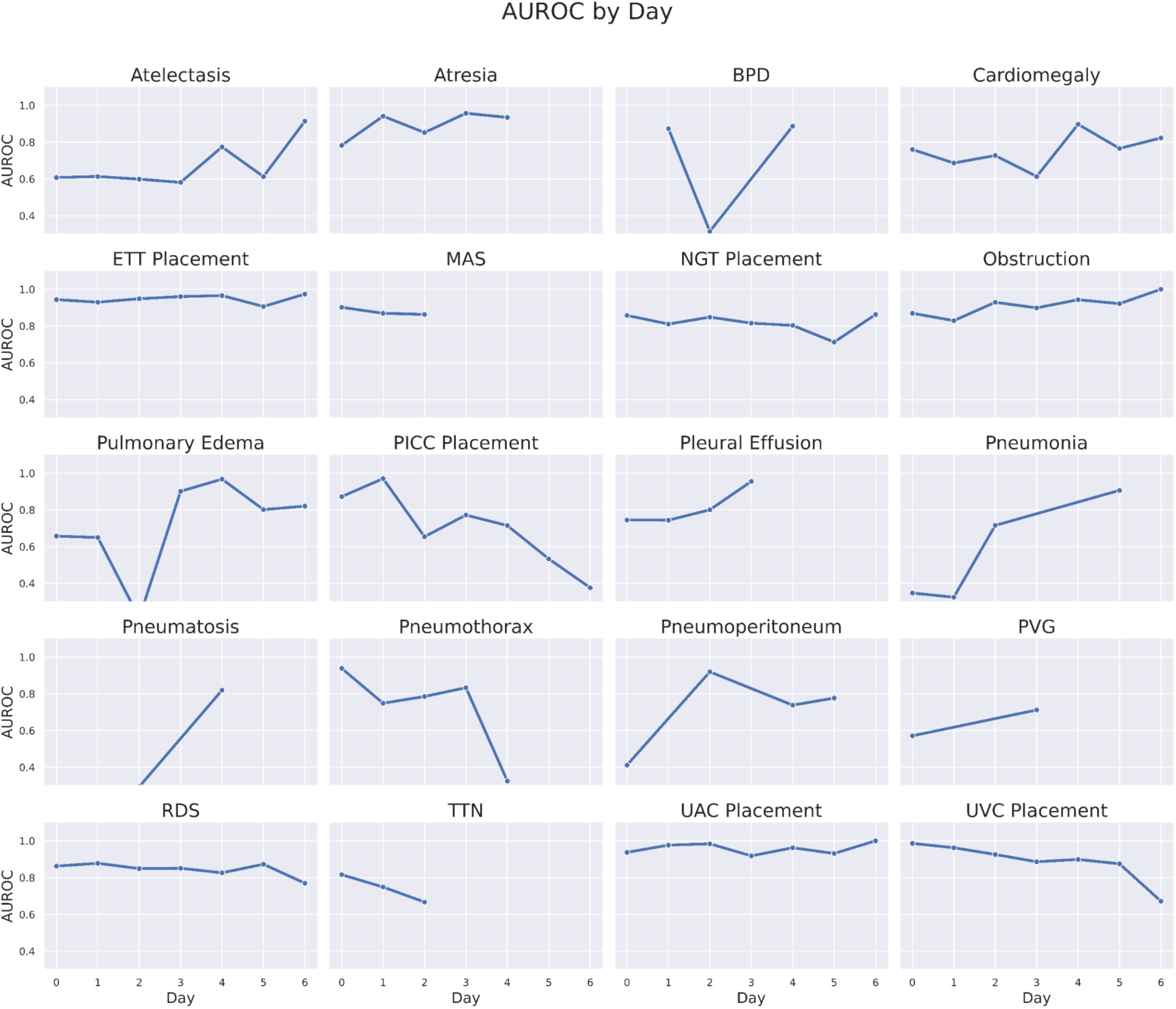
AUROC of Our Model by the Day Image Was Obtained During First Week of Life.

**Figure 3.**
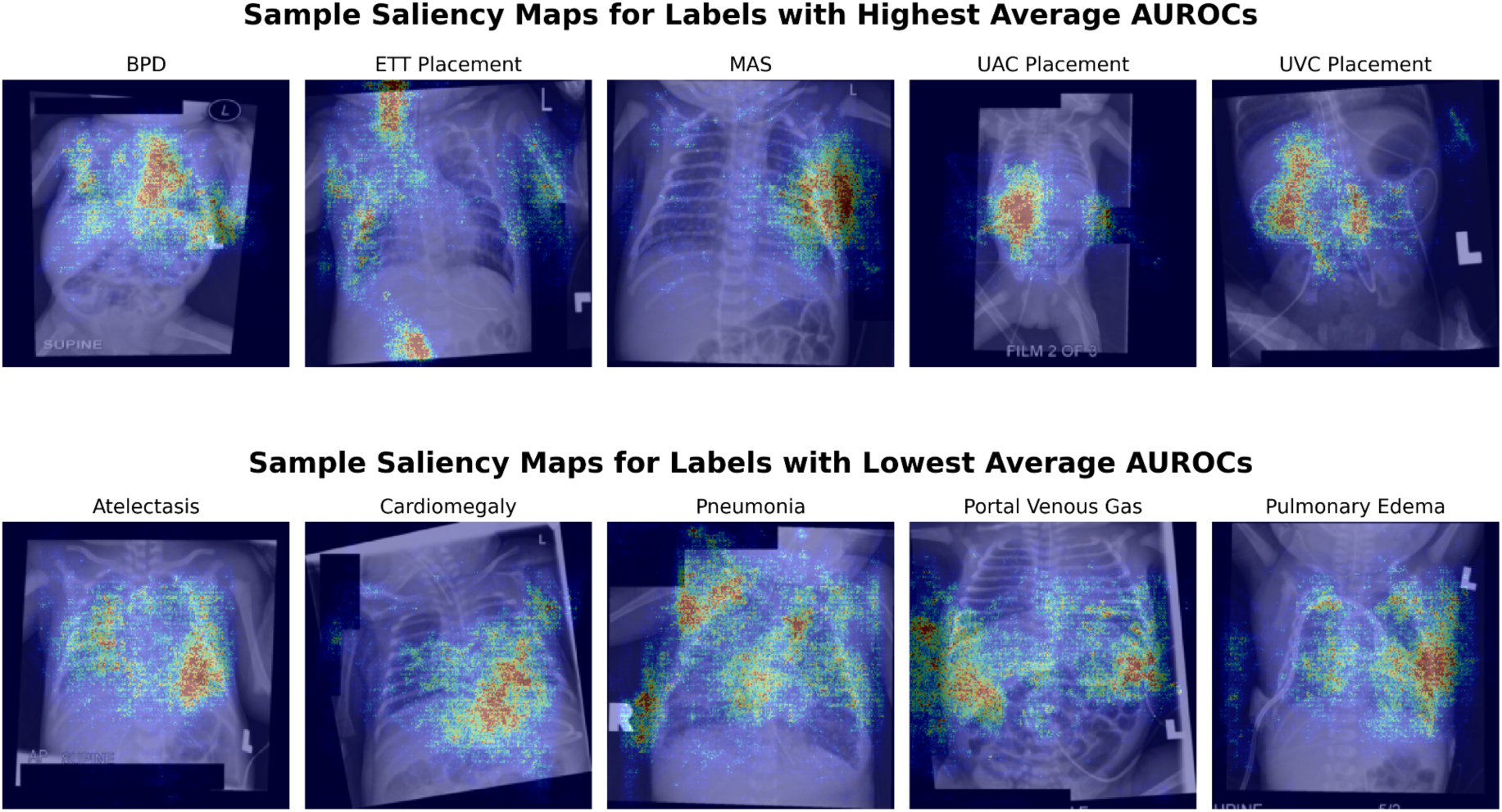
Sample Saliency Maps for the Best and Worst Performing Labels in Our Model.

## Discussion

With this large cohort of infants, we developed a novel CLIP-based foundation model which accurately interpreted neonatal radiographs to identify numerous pathologies and findings relevant to neonatal intensive care. This model, NeoCLIP, outperformed logistic regression, ResNet-50, and BioViL-T in all of the labels except for intestinal atresia, MAS, and portal venous gas, although the differences for these three labels are not statistically significant. While the addition of gestational age and birthweight did improve the performance of our novel model, it was not statistically significant in the majority of labels.

This study marks the first application of deep contrastive learning techniques on radiograph-report pair pre-training for interpreting neonatal radiographs in a cohort of infants admitted to the NICU. Our model is able to identify common neonatal pathologies and medical devices on radiographs with great success. The performance of NeoCLIP is similar to models trained on and applied to adult and pediatric populations (21-24). It is noteworthy, however, that our model was tasked with identifying more distinct labels than these similar adult and pediatric studies. We also compared the performance of NeoCLIP to other imaging models on our cohort of radiographs. NeoCLIP outperformed ResNet-50 on all labels except intestinal atresia. While it is unclear why ResNet-50 performed the best for intestinal atresia, it may be due to the small number of positive cases (**Table 1**) as well as the LLM labeler having the lowest accuracy on this label (**Table 2**). For all other labels, given that NeoCLIP was pre-trained and fine-tuned on our neonatal radiographs, its improved performance is not unexpected when compared to a model trained on a generalized database such as ResNet-50. Similarly, our model outperforms BioViL-T in nearly all labels other than portal venous gas, which also has a small number of positive cases (**Table 1**). Much like ResNet-50, BioViL-T is not pre-trained on neonatal images, rather it is pre-trained on databases of adult radiographs (15). The improved performance of our model highlights the importance of pre-training and potentially fine-tuning a model on neonatal radiographs. This notion is further reinforced by the fact that BioViL-T does not seem to outperform ResNet-50 consistently, despite the fact that it is pre-trained on adult radiographs. This suggests that a model pre-trained on medical images will not perform well on similar studies in a different population, highlighting the importance of training models for specific demographics. As such, novel investigations such as ours are essential to ensuring that neonates get similar quality of care to other populations.

Our investigation also reinforces the importance of gestational age and birth weight as clinical risk factors for many neonatal pathologies (25-28). While NeoCLIP significantly outperformed the logistic regression based on just the two demographic factors alone (**Table 3**), the performance of our model was boosted by addition of gestational age and birth weight. While this improvement is not always statistically significant, it does further stress the importance of these clinical factors and therefore highlight the potential importance of a multimodal model even with image-related tasks. We also noted that NeoCLIP performs better on the labels related to medical devices than those related to pathology. This was expected given that the presence of these devices is a more specific outcome than many of the pathologic labels. This is further emphasized by the worse performance of NeoCLIP on subjective findings such as cardiomegaly and atelectasis when compared to well-defined radiographic findings such as pneumothorax or pneumatosis. It is worth noting, however, that our model did not identify correct placement of any medical devices, only whether they were present. An interesting future study would be to assess the model’s performance in identifying the correct placement of these critical lines and tubes.

Furthermore, we found that the performance of NeoCLIP is relatively stable as a function of time. When stratifying the images by week, we found the AUROCs remain consistent for the majority of labels (**Figure 1**). There did appear, however, to be less stability of AUROCs when stratified by day within the first week of life (**Figure 2**). This discrepancy is likely more of a function of positive sample size than inherent variability in the model’s performance. Namely, there were likely less images captured in a specific day than a week. While the stability of the model is reassuring, it is important to assess those labels where the model does not seem to have consistency. For example, the labels of pneumonia and UVC placement provide important insight into the model. Pneumonia is typically diagnosed after the first few days of life; this is reflected by our model in that the AUROC becomes much more stable after day 3. Similarly, the AUROC on the weekly level increases overall by weeks 5-7 indicating that more infants may be included in that time period with a diagnosis of pneumonia. Similarly, UVCs are usually discontinued by day 10, which is consistent with the observed decrease in the previously stable AUROCs for UVC placement. This time series data allows us to interpret how a model’s performance may change as more or less data becomes available at a given time point.

To interrogate the model and investigate what signals may be contributing to the predicted diagnoses, we created saliency maps (**Figure 3**). The red areas of the saliency maps indicate what areas the model is paying the most attention to versus the blue areas where the model is paying least attention. For the best performing labels, we see that the concentration of red is over the anatomically and/or physiologically accurate locations. For both ETT and UAC placement, we see a high concentration of red over where these specific devices would appear on a radiograph. For BPD and MAS, the model is appropriately focused on the lung regions of the radiograph. In addition to this accuracy, the model appears to also be precise in that there are not multiple areas on the radiographs where there is a concentration of red. For the worst performing labels, while accurate, there seems to be less precision. The focus of the model is consistent with the labels - cardiomegaly shows concentration around the heart, portal venous gas around the liver, and the pulmonary findings in the lungs. However, in direct contrast with the best performing labels, there are multiple concentrations of red showing a lack of precision. This ambiguity is likely a function of the labels as many of these diagnoses, such as pulmonary edema or atelectasis, represent a diffuse radiologic process. While saliency maps do not provide complete insight into a neural network, they do allow us to make inferences that can help us better interpret and explain the performance of a model. As such, these maps will remain valuable tools as deep learning becomes increasingly embedded in clinical practice.

Another key finding of our investigation was the innovative application of a LLM to generate labels from radiograph reports. Not every corresponding report to a radiograph in our cohort included a label consistent with those of interest for our investigation. As such, we prompted a LLM to identify disease and device labels for a particular radiograph based on the text of the report. The accuracy, sensitivity, and specificity of the LLM in identifying correct labels for a subgroup of imaging demonstrated this technique as a proof of concept. To our knowledge, this methodology has not been employed prior in the neonatal population prior to our investigation and therefore represents a potentially impactful technique for extracting labels for unlabeled images or data when training and validating complex deep learning models.

Despite the strengths of this study, it is not without limitations. This model was developed using single center data in addition to being internally validated on the same cohort it was trained on. While this is a common methodological approach, it does limit the generalizability of the study. An interesting next step would be to apply our novel on a unique cohort of infants admitted to geographically distinct NICU. Furthermore, there remains an inherent issue with explainability that is consistent with all deep learning studies and not unique to our investigation. While the saliency maps do provide some insight into the decision-making process of the model, it still does not fully explain what aspects of the image factor into its task. Another limitation of our study is the heterogeneous nature of our cohort when it comes to gestational age. We did not exclude any infants based on gestational age and therefore the mean gestational age of our cohort at birth was roughly 34 weeks (**Table 1**). However, many of the pathologies we asked the model to identify are unique to gestational ages - such as RDS or BPD for preterm infants and TTN for term infants. An interesting next step would be to train a model specific to gestational age for these specific pathologies. We did not pursue this as we wanted to maximize the number of radiographs for training the model and felt any gestational age limitation would hinder our ability to adequately train our model. Despite this approach, there still remains small sample sizes for certain labels such as intestinal atresia, pneumoperitoneum, and portal venous gas (**Table 1**). While the concept of statistical power is not always consistent with deep learning studies, the lower sample sizes do challenge the validity of some of our results. Additionally, there is inherent bias due to the retrospective nature of our study. All radiographs were obtained due to a clinical indication, and therefore the very presence of a radiograph suggests some level of provider concern for a certain disease process. However, this methodological bias could only be avoided by taking radiographs of all infants which is not feasible due to the potential harm of radiation. Despite these limitations, this investigation remains one of the largest such studies of this type in the neonates. Its novel methodology and promising results represent a major step in the application of deep learning principles to this vulnerable population.

## Conclusions

In this large retrospective study, we trained and validated a novel, deep learning model which successfully identified a number of common pathologies and medical devices on neonatal radiographs. Our model, NeoCLIP, outperforms similar models trained and developed on adult populations. This represents the first such application of advanced machine learning methodologies to interpret neonatal radiographs. Furthermore, our methodology represents the first application of a LLM to extract disease state labels from neonatal radiology reports to fine tune an imaging model. While NeoCLIP effectively interprets neonatal radiographs, it is not without limitations in generalizability and interpretability. Despite these limitations, however, our investigation represents a major step in the application of artificial intelligence to interpret medical data to diagnose disease states and impact clinical care.

## Data Availability

All data produced in the present study are available upon reasonable request to the authors.

## Author Contributions

KB and AB conceived of the project. KB, AB, and NG assisted in enrollment of the cohort. KB, AB, NG, YH, PS, and AP participated in data analysis. YH, PS, AP, and KB drafted the initial manuscript. All authors reviewed and approved of final submission.

## Acknowledgements

The authors would like to acknowledge all the patients and families whose data was used in the training and validation of this model.

## Data Statement

All data generated or analyzed during this study are included in this published article (and its supplementary information files).

## References

1. Gopal G, Suter-Crazzolara C, Toldo L, et al. Digital transformation in healthcare – architectures of present and future information technologies. Clin Chem Lab Med. 2019; 57(3): 328–335. doi:10.1515/cclm-2018-0658.

2. Beam K, Sharma P, Levy P, et al. Artificial intelligence in the neonatal intensive care unit: the time is now. J Perinatol. 2024;44(1):131–135. doi:10.1038/s41372-023-01719-z.

3. McBee MP, Awan OA, Colucci AT, et al. Deep Learning in Radiology. Acad Radio. 2018;25:1472–1480. doi:10.1016/j.acra.2018.02.018.

4. Dillman JR, Somasundaram E, Brady SL, et al. Current and emerging artificial intelligence applications for pediatric abdominal imaging. Pediatr Radiol. 2022;52(11):2139–2148. doi:10.1007/s00247-021-05057-0.

5. Saba L, Biswas M, Kuppili V, et al. The present and future of deep learning in radiology. Eur J Radiol. 2019 May;114:14–24. doi:10.1016/j.ejrad.2019.02.038.

6. Chen T, Kornblith S, Norouzi M, et al. A Simple Framework for Contrastive Learning of Visual Representations. In Proceedings of the 37th International Conference on Machine Learning. 2020;119(149):1597-1607.

7. Tiu E, Talius E, Patel P, et al. Expert-level detection of pathologies from unannotated chest X-ray images via self-supervised learning. Nat Biomed Eng. 2022;6(12):1399–1406. doi:10.1038/s41551-022-00936-9.

8. Wu C, Zhang X, Zhang Y, et al. Medical knowledge enhanced language-image pre-training for x-ray diagnosis. In Proceedings of the IEEE/CVF International Conference on Computer Vision. 2023:21372–21383. doi:10.1109/ICCV51070.2023.01954

9. Moor M, Banerjee O, Abad ZSH, et al. Foundation models for generalist medical artificial intelligence. Nature. 2023;616(7956):259-265. doi:10.1038/s41586-023-05881-4.

10. Palepu A, Beam A. Tier: Text-image entropy regularization for medical clip-style models. In Proceedings of the 8th Machine Learning for Healthcare Conference. 2023;219:548-564.

11. Hysinger EB, Higano NS, Critser PJ, et al. Imaging in neonatal respiratory disease. Paediatr Respir Rev. 2022;43:44–52. doi:10.1016/j.prrv.2021.12.002.

12. Liu J. Ultrasound diagnosis and grading criteria of neonatal respiratory distress syndrome. J Matern Fetal Neonatal Med. 2023;36(1):2206943. doi:10.1080/14767058.2023.2206943.

13. Rich BS, Dolgin SE. Necrotizing Enterocolitis. Pediatr Rev. 2017;38(12):552-559. doi:10.1542/pir.2017-0002.

14. Gore JC. Artificial intelligence in medical imaging. Magn Reson Imaging. 2020;68:A1–A4. doi:10.1016/j.mri.2019.12.006.

15. Bannur S, Hyland S, Liu Q, et al. Learning to exploit temporal structure for biomedical vision-language processing. arXiv.org. Published March 16, 2023. doi:10.48550/arXiv.2301.04558.

16. Kenton JD, Chang MW, Toutanova K. BERT: Pre-training of deep bidirectional transformers for language understanding. In Proceedings of NAACL-HLT. Vol 1. 2019:2.

17. He K, Zhang X, Ren S, et al. Deep Residual Learning for Image Recognition. In IEEE Conference on Computer Vision and Pattern Recognition. 2016: 770–8.

18. Radford A, Kim JW, Hallacy C, et al. Learning transferable visual models from natural language supervision. In International Conference on Machine Learning. PMLR; 2021:8748–8763.

19. Krizhevsky A, Sutskever I, Hinton GE. ImageNet classification with deep convolutional neural networks. In Advances in Neural Information Processing Systems. 2012:1097–1105. doi:10.1145/3065386.

20. Shorten C, Khoshgoftaar TM. A survey on image data augmentation for deep learning. J Big Data. 2019;6(1):1–48. doi:10.1186/s40537-019-0197-0.

21. Ahn JS, Ebrahimian S, McDermott S, et al. Association of Artificial Intelligence-Aided Chest Radiograph Interpretation With Reader Performance and Efficiency. JAMA Netw Open. 2022;5(8):e2229289. doi:10.1001/jamanetworkopen.2022.29289.

22. Bennani S, Regnard NE, Ventre J, et al. Using AI to Improve Radiologist Performance in Detection of Abnormalities on Chest Radiographs. Radiology. 2023;309(3):e230860. doi:10.1148/radiol.230860.

23. Padash S, Mohebbian MR, Adams SJ, et al. Pediatric chest radiograph interpretation: how far has artificial intelligence come? A systematic literature review. Pediatr Radiol. 2022;52(8):1568–1580. doi:10.1007/s00247-022-05368-w.

24. Lind Plesner L, Müller FC, Brejnebøl MW, et al. Commercially Available Chest Radiograph AI Tools for Detecting Airspace Disease, Pneumothorax, and Pleural Effusion. Radiology. 2023;308(3):e231236. doi:10.1148/radiol.231236.

25. Jang W, Choi YS, Kim JY, et al. Artificial Intelligence-Driven Respiratory Distress Syndrome Prediction for Very Low Birth Weight Infants: Korean Multicenter Prospective Cohort Study. J Med Internet Res. 2023;25:e47612. doi:10.2196/47612.

26. Condò V, Cipriani S, Colnaghi M, et al. Neonatal respiratory distress syndrome: are risk factors the same in preterm and term infants? J Matern Fetal Neonatal Med. 2017;30(11):1267–1272. doi:10.1080/14767058.2016.1210597.

27. Jung E, Lee BS. Late-Onset Sepsis as a Risk Factor for Bronchopulmonary Dysplasia in Extremely Low Birth Weight Infants: A Nationwide Cohort Study. Sci Rep. 2019;9(1):15448. doi:10.1038/s41598-019-51617-8.

28. Gibbs K, Jensen EA, Alexiou S, et al. Ventilation Strategies in Severe Bronchopulmonary Dysplasia. Neoreviews. 2020;21(4):e226–e237. doi:10.1542/neo.21-4-e226.

